# A Novel Dual-Outcome Risk Score for VBAC Success and Neonatal Morbidity

**DOI:** 10.64898/2026.07.01.26357032

**Authors:** Lauren Crabtree, Ruofan Yao, Ciprian P. Gheorghe

## Abstract

**Objective:** To develop and externally validate a simple antepartum cumulative risk score that stratifies both vaginal birth after cesarean (VBAC) success and neonatal morbidity among patients undergoing trial of labor after cesarean (TOLAC).

**Methods:** This retrospective cohort study was conducted in 2 stages: model development in a single tertiary care center in California (2019–2025) and external validation in the National Vital Statistics System natality files (2020–2024). The derivation cohort included 1,418 TOLAC attempts; the national validation cohort included 477,693 TOLAC attempts. A point-based score was constructed from routinely available antepartum characteristics associated with VBAC. VBAC success and neonatal intensive care unit (NICU) admission were evaluated across score levels in both cohorts, and model discrimination was assessed using area under the receiver operating characteristic curve (AUC).

**Results:** In the derivation cohort, 1,087 of 1,418 patients (76.7%) achieved VBAC. The logistic regression model showed reasonable discrimination (AUC 0.70, 95% CI 0.67–0.73). VBAC success declined from 89.1% at a score of −1 to 37.8% at scores of 4 or higher, whereas NICU admission increased from 31.7 to 200.0 per 1,000. Uterine rupture occurred in 28 of 1,418 TOLAC attempts (1.97%) and was not predicted by antepartum characteristics. In the national cohort, VBAC success similarly declined from 90.5% to 44.8%, whereas NICU admission increased from 43.8 to 111.1 per 1,000 across the same score range.

**Conclusion:** A simple antepartum risk score stratified both VBAC success and neonatal morbidity in single-center and national TOLAC cohorts, supporting its potential use in patient-centered counseling.

## INTRODUCTION

Trial of labor after cesarean (TOLAC) is a crucial shared decision in obstetric practice. When successful, vaginal birth after cesarean (VBAC) reduces morbidity in future pregnancies; when unsuccessful, failed TOLAC confers risk that exceeds that of elective repeat cesarean.^1-3^ In the United States, cesarean delivery now accounts for more than one in three births, creating a growing population of patients for whom subsequent delivery planning centers on the decision to pursue TOLAC.^4^ Pregnancy and delivery complications contribute substantial health care and societal costs, underscoring the importance of reducing preventable morbidity where safe alternatives to surgery exist.^5-7^ From a public health perspective, optimizing VBAC among appropriate candidates is a mechanism not only for improving maternal outcomes but also for mitigating the downstream costs of repeat operative delivery at scale.

Despite guideline support for TOLAC, access in the United States remains limited and geographically uneven.^8-10^ Hospitals offering TOLAC are clustered in a minority of US counties, with large regions of the country lacking local access, constraining the potential population-level benefits of TOLAC and contributing to variation in VBAC rates unlikely to be explained by patient risk alone.^9-10^ There is therefore a need for tools that use routinely available clinical variables to provide clear estimates of VBAC success and neonatal morbidity and can be used in busy clinical settings to support patient-centered counseling about TOLAC.

Several TOLAC prediction models exist, including the widely used Grobman model duderived from the Maternal-Fetal Medicine Units Network (MFMU).^11-12^ However, most population-based VBAC calculators were developed more than two decades ago in cohorts that predated the increases in maternal age, obesity, and medical comorbidities that characterize contemporary obstetric populations.^11-14^ In the current era, these models may misclassify risk for many TOLAC candidates, particularly those with more complex health profiles. Further, existing VBAC calculators often provide broad probability ranges that limit the precision of individual counseling, reduce their clinical applicability, and offer limited leverage for health systems to standardize counseling and expand access to TOLAC at a population level. More complex modeling approaches, including machine-learning methods, have been explored, but have not produced widely-used, clinically simple tools for TOLAC counseling.^15^

A further limitation of existing models is the singular focus on VBAC success as the outcome of interest. From the perspective of patient counseling, the relevant question is not merely whether vaginal delivery will be achieved, but whether the overall maternal and neonatal outcome profile favors TOLAC for a given patient. Neonatal morbidity represents a clinically meaningful endpoint that has not been incorporated into VBAC prediction frameworks or TOLAC decision aids. Incorporating neonatal outcomes alongside maternal risks is particularly important in contemporary obstetric populations with higher baseline risk profiles, where small differences in intrapartum management may substantially affect neonatal resource use and early-life morbidity. A tool that provides clear estimates of both VBAC success and neonatal morbidity using routinely available clinical information could support patient-centered counseling and inform system-level efforts to expand access to TOLAC.

In this context, we sought to develop a simple cumulative risk score, using routinely available clinical characteristics, that stratifies both VBAC success and neonatal morbidity among patients undergoing TOLAC and to assess the extent to which uterine rupture can be predicted from antepartum factors. We then evaluated whether this dual-outcome TOLAC risk score retains its performance in contemporary US practice by applying it to a national sample of term singleton TOLAC deliveries.

## METHODS

### Study Design

We performed a two-stage retrospective cohort study: model development in an institutional TOLAC cohort and external validation in a national TOLAC cohort. Model development used TOLAC data from a tertiary care center in California (April 2019-December 2025). External validation used term singleton TOLAC data from United States deliveries (2020-2024), as recorded in the Centers for Disease Control and Prevention (CDC) National Vital Statistics System (NVSS) natality files. Institutional review board approval was obtained for the institutional cohort (IRB#5260029); IRB review was not required for the national cohort because the NVSS data are publicly available and fully deidentified.

### Derivation Cohort

All patients who underwent TOLAC at a single tertiary care center in California during the study period were identified from an institutional perinatal quality database. The dataset included 1,419 consecutive TOLAC cases; after exclusion of one case with a biologically implausible BMI, the final analytic derivation cohort comprised 1,418 TOLAC attempts. All analyses in the derivation cohort used complete-case data.

#### Outcomes

The primary outcome was successful VBAC, defined as vaginal delivery (spontaneous, vacuum-assisted, or forceps-assisted) after a prior cesarean. Secondary outcomes were neonatal intensive care unit (NICU) admission, severe maternal morbidity (SMM, excluding transfusion-only cases per Centers for Disease Control and Prevention criteria), and uterine rupture identified by International Classification of Diseases, Tenth Revision (ICD-10) code O71.1 (rupture of uterus during labor) within the composite diagnosis field.

#### Predictor Variables

Candidate predictors at delivery counseling were maternal age, pre-pregnancy BMI, gestational age, parity, race and ethnicity (self-reported), labor onset (spontaneous vs. induced), diabetes (none, gestational, pregestational), hypertensive disorders (none, chronic hypertension, preeclampsia–eclampsia), premature rupture of membranes (PROM), chorioamnionitis, and prior vaginal delivery. Prior vaginal delivery was inferred from parity greater than 1, consistent with local practice that patients with more than one prior cesarean are not offered TOLAC.

#### Model Development and Internal Validation

Bivariable associations between candidate predictors and VBAC were assessed using chi-square or Fisher exact tests for categorical variables and Mann–Whitney U tests for continuous variables with nonnormal distributions. Multivariable logistic regression with backward elimination (*P* < 0.10) was used to develop a prediction model for VBAC, and results were reported as adjusted odds ratios with 95% confidence intervals. Model discrimination was summarized using the area under the receiver operating characteristic curve, and calibration was evaluated with the Hosmer-Lemeshow goodness-of-fit test and calibration plots. In parallel, four machine learning algorithms (penalized logistic regression with L2 regularization, random forest, gradient boosting, and extreme gradient boosting [XGBoost]) were trained with 5-fold stratified cross-validation, with hyperparameters selected to maximize cross-validated area under the curve; feature importance was derived from the random forest model, and calibration for all models was examined using calibration plots.

### Cumulative Risk Score

A clinically-applicable risk score was developed from independent predictors identified in the multivariate logistic regression and machine learning, weighted by the magnitude and direction of their associations. Points were assigned as follows: pre-pregnancy body mass index (BMI) 30 or greater (+1 point), BMI 40 or greater (+1 additional point), induction of labor (IOL; +1), diabetes mellitus (pregestational or gestational; +1), hypertensive disorder (chronic hypertension, gestational hypertension, or eclampsia; +1), maternal age 40 years or older (+1), and gestational age 41 weeks or greater (+1). Prior vaginal delivery was assigned −1 point, yielding a total score range of −1 to 7. VBAC success rates and NICU admission rates were calculated at each score level with 95% confidence intervals, and the association between increasing risk score and these outcomes was assessed using Spearman rank correlation.

### National Validation Cohort

The external validation cohort included patients who underwent TOLAC among term singleton deliveries in the United States from 2020 through 2024 in the NVSS natality files. TOLAC was identified using two criteria applied in combination: the presence of at least one prior cesarean delivery (recorded in the risk factor checkbox on the birth certificate) and evidence of a trial of labor, defined as either a vaginal delivery after prior cesarean (successful VBAC) or a cesarean delivery with the trial of labor field marked as attempted (failed TOLAC). Patients who delivered by repeat cesarean without a documented TOLAC were classified as elective repeat cesarean and excluded. The cohort was restricted to term (37-42 weeks) singleton deliveries, yielding a final national TOLAC cohort of 477,693 attempts. Analyses of the national cohort used complete-case data for the variables included in each model.

#### Outcomes

The primary outcome in the national cohort was successful VBAC, using the same definition as in the derivation cohort. The secondary outcome was NICU admission, as recorded on the birth certificate. Uterine rupture could not be assessed because it is not captured in the NVSS natality files.

#### Application of the Cumulative Score

In the national NVSS cohort, the cumulative risk score was calculated for each term singleton TOLAC attempt using the same point assignments defined in the derivation cohort. VBAC success and NICU admission were summarized across score levels to evaluate whether the score maintained its discrimination in the national population.

### Statistical Analysis

Analytic methods for model development, internal validation, and cumulative risk score performance in the single-center derivation cohort are described in previous sections. In the national NVSS cohort, associations between individual risk factors and VBAC success were expressed as absolute differences in VBAC rates, the crude association between induction of labor and failed TOLAC was estimated as an odds ratio with 95% confidence intervals, predictors of VBAC success were evaluated with logistic regression and receiver operating characteristic curves, and VBAC rates and TOLAC volume were examined by year.

All analyses were conducted using Python version 3.10 with NumPy, pandas, SciPy, statsmodels, scikit-learn, and XGBoost. A two-tailed P value less than 0.05 was considered statistically significant, with random seeds fixed at 42 to support reproducibility. Anthropic Claude (Anthropic) was used to assist with generation of Python code for data extraction and statistical analyses; all code output was reviewed, verified, and edited by the authors.

## RESULTS

### Derivation Cohort

#### Cohort Characteristics

A total of 1,418 patients attempted TOLAC at the single tertiary care center. Median maternal age was 31 years, median pre-pregnancy BMI was 28.4 kg/m^2^ (24.8–33.3), and median gestational age at delivery was 39.1 weeks (38.4–40.0). The cohort was predominantly Hispanic (63.1%), followed by White (15.8%), Black (8.7%), multiracial (5.3%), Asian (4.0%), and other (3.1%). Labor was induced in 43.4% of cases; 12.6% had diabetes mellitus, 24.1% had hypertensive disorders, 6.6% had premature rupture of membranes, 7.3% developed chorioamnionitis, and 58.3% had a prior vaginal delivery (Table 1).

**Table 1.** Demographic and Clinical Characteristics of the Single-Center Population.

| Variable | Overall<br>(N=1,418) | VBAC Success<br>(n=1087) | Failed TOLAC<br>(n=331) | P value |
| --- | --- | --- | --- | --- |
| Age (SD) | 31.1 (5.5) | 31.0 (5.5) | 31.3 (5.5) | 0.3594 |
| BMI (SD) | 29.5 (6.6) | 28.9 (6.4) | 31.2 (7.0) | <.001 |
| Gestational Age (SD) | 39.2 (1.7) | 39.1 (1.8) | 39.4 (1.2) | 0.0039 |
| Induction of Labor | 615 (43.4%) | 418 (38.5%) | 197 (59.5%) | <.001 |
| Diabetes Mellitus | 178 (12.6%) | 120 (11.0%) | 58 (17.5%) | 0.0025 |
| Hypertensive Disorder | 342 (24.1%) | 227 (20.9%) | 115 (34.7%) | <.001 |
| Obesity (BMI ≥ 30) | 569 (40.1%) | 403 (37.1%) | 166 (50.2%) | <.001 |
| Prior Vaginal Delivery | 826 (58.3%) | 693 (63.8%) | 133 (40.2%) | <.001 |
| Age ≥ 40 years | 93 (6.6%) | 68 (6.3%) | 25 (7.6%) | 0.4790 |
| GA ≥ 41 weeks | 96 (6.8%) | 60 (5.5%) | 36 (10.9%) | 0.0011 |
| PROM | 93 (6.6%) | 68 (6.3%) | 25 (7.6%) | 0.4790 |
| **Race/Ethnicity** |  |  |  |  |
| Hispanic US Born | 621 (43.8%) | 480 (44.2%) | 141 (42.6%) |  |
| Hispanic Non-US Born | 273 (19.3%) | 219 (20.1%) | 54 (16.3%) |  |
| White | 224 (15.8%) | 172 (15.8%) | 52 (15.7%) |  |
| Black | 123 (8.7%) | 87 (8.0%) | 36 (10.9%) |  |
| Multiracial | 75 (5.3%) | 50 (4.6%) | 25 (7.6%) |  |
| Asian | 57 (4.0%) | 48 (4.4%) | 9 (2.7%) |  |
| Unknown | 29 (2.0%) | 21 (1.9%) | 8 (2.4%) |  |
| Pacific Islander | 11 (0.8%) | 7 (0.6%) | 4 (1.2%) |  |
| Native American | 4 (0.3%) | 2 (0.2%) | 2 (0.6%) |  |
| Other | 1 (0.1%) | 1 (0.1%) | 0 (0.0%) |  |
| **Outcomes** |  |  |  |  |
| NICU admission | 103 (7.3%) | 57 (5.2%) | 46 (13.9%) |  |
| Uterine rupture | 28 (2.0%) | 3 (0.3%) | 25 (7.6%) |  |
| Severe maternal morbidity | 13 (0.9%) | 5 (0.5%) | 8 (2.4%) |  |

#### VBAC Prediction and Cumulative Score Performance

Of the 1,418 patients who attempted TOLAC, 1,087 (76.7%) had a successful VBAC; this rate was consistent across study years. The logistic regression model used to build the score showed reasonable discrimination (AUC 0.70, 95% CI 0.6–0.73) with good calibration, and a penalized logistic model achieved a nearly identical cross-validated AUC of 0.71 (95% CI 0.67– 0.75). As scores increased, VBAC success declined from 89.1% at a score of −1 to 37.8% at scores of 4 or higher (P < .01; Figure 1), while NICU admission rose from 31.7 to 200.0 per 1,000 (P < .01; Figure 2).

**Figure 1.**
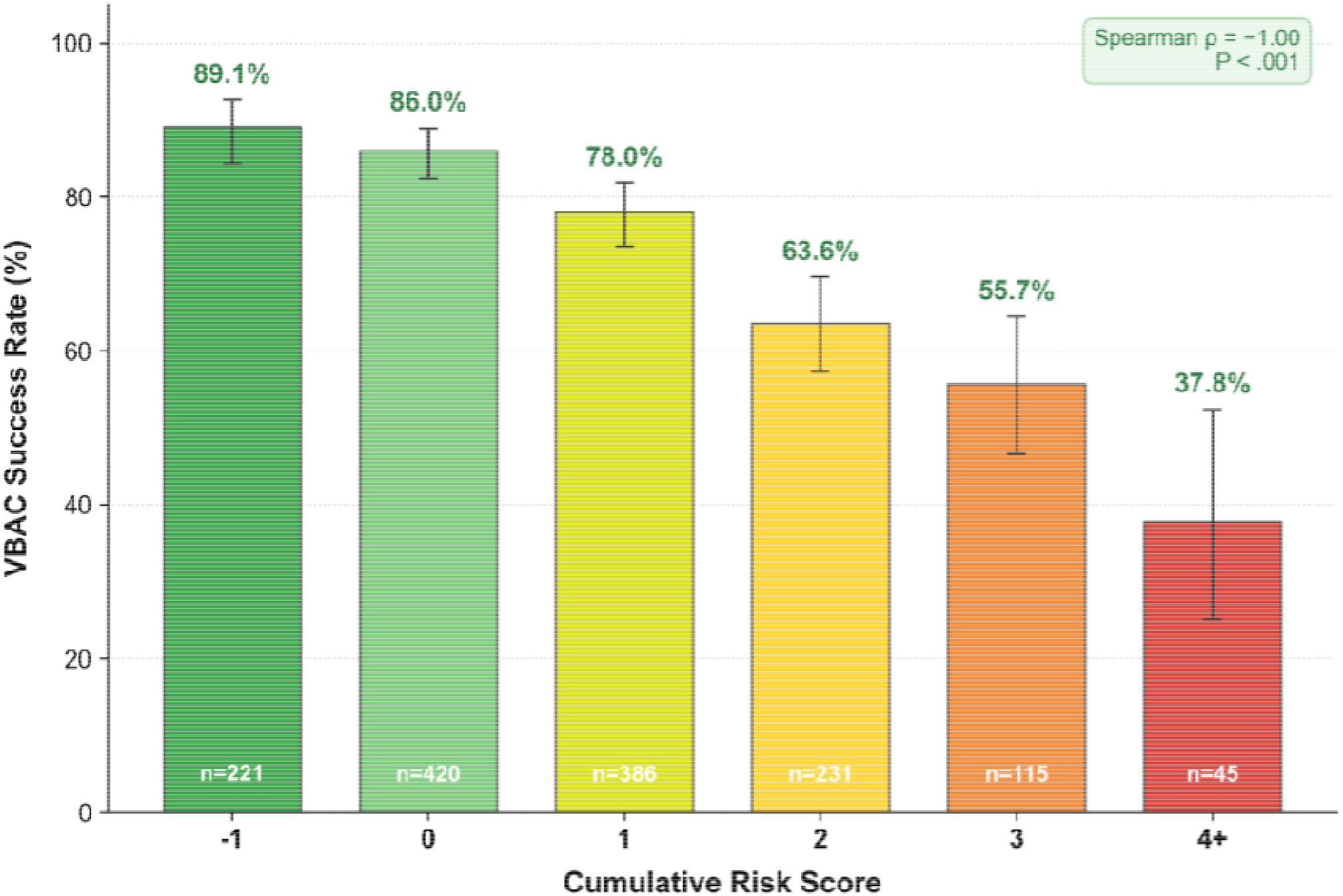
VBAC success rate by cumulative risk score in the single-center derivation cohort. Bars show the percentage of patients achieving VBAC at each cumulative risk score level (−1, 0, 1, 2, 3, and ≥4), with 95% confidence intervals indicated by error bars and the number of TOLAC attempts at each score displayed within bars.

**Figure 2.**
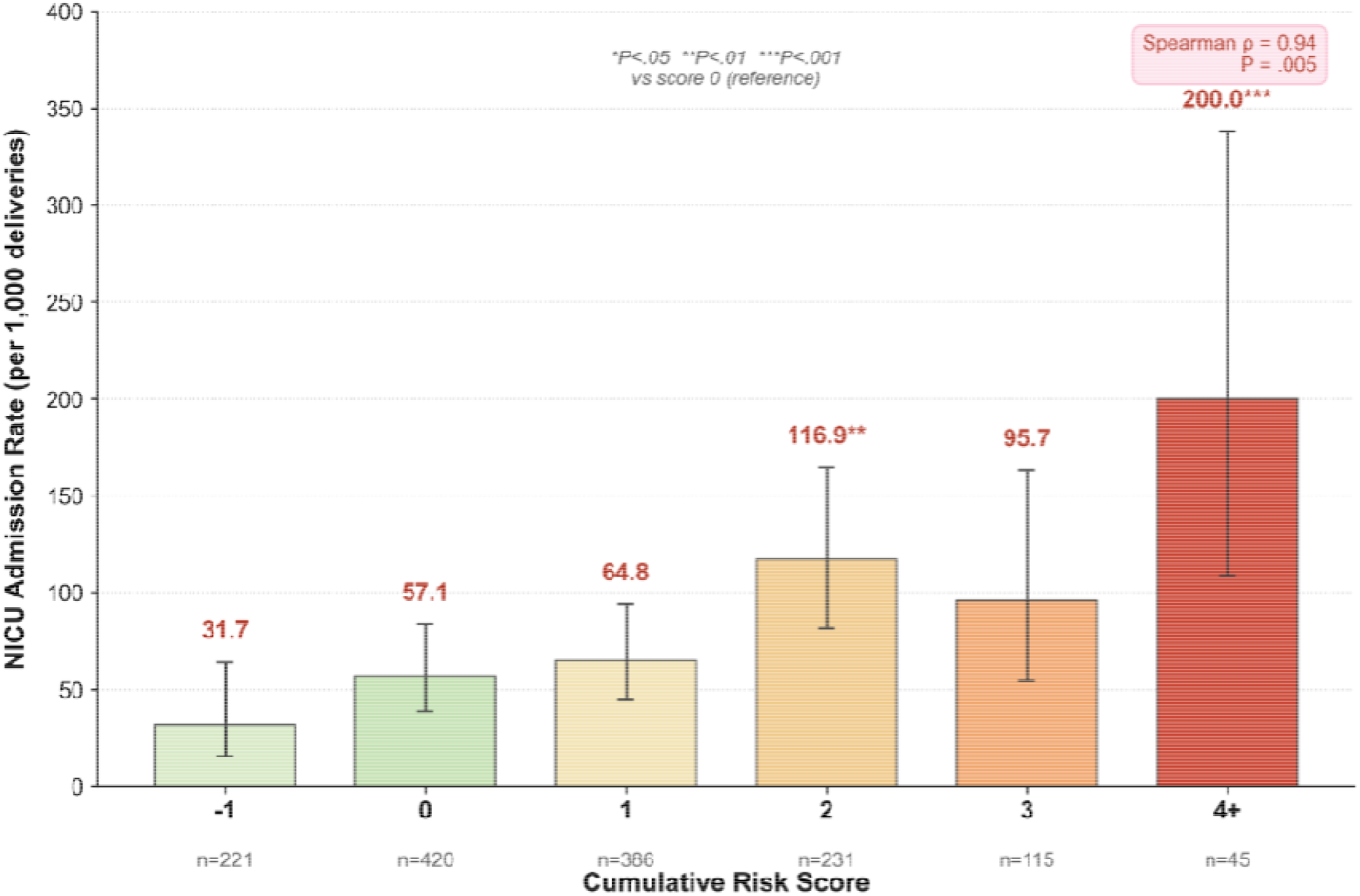
NICU admission rate by cumulative risk score in the single-center derivation cohort. Bars show NICU admission rates per 1,000 deliveries at each cumulative risk score level (−1, 0, 1, 2, 3, and ≥4), with 95% confidence intervals indicated by error bars and the number of TOLAC attempts at each score displayed within bars.

#### Uterine Rupture and Severe Maternal Morbidity

Uterine rupture occurred in 28 of 1,418 TOLAC attempts (1.97%). Severe maternal morbidity was more common among patients with rupture than those without (10.7% vs 0.7%; odds ratio 16.56, 95% CI 4.32–63.42, P < .001). No maternal or antepartum characteristic, including the cumulative risk score, was significantly associated with uterine rupture.

#### Exclusion Threshold Analysis

Excluding patients with risk scores of 3 or higher (n=160, 11.3% of the cohort) increased overall VBAC success from 76.7% to 80.0% (P=.04) and reduced NICU admission from 72.6 to 66.0 per 1,000 (P=.50). A more stringent exclusion of scores 2 or higher (n=391, 27.6%) increased VBAC success to 83.6% (P<.001) and reduced NICU admission to 54.5 per 1,000 (P=.07). Uterine rupture rates were not significantly altered by either threshold (P=.94 and P=.87, respectively). Among the 160 excluded patients (score 3 or higher), the observed VBAC rate was 50.6% and the NICU admission rate was 125.0 per 1,000, both significantly different from the retained cohort (P<.001 and P=.007, respectively).

### National Validation Cohort

#### Cohort Characteristics

A total of 477,693 patients met TOLAC inclusion criteria in the NVSS natality files from 2020 through 2024. Mean maternal age was 31 years, mean pre-pregnancy BMI was 28, and mean gestational age at delivery was 39 weeks. Labor was induced in 29.6% of cases; 9.7% had diabetes mellitus, 9.8% had hypertensive disorders, and 45.4% had a prior vaginal delivery. The overall VBAC rate in the national cohort was 73.3% (Table 2).

**Table 2.** Cohort Characteristics of the National Cohort.

| Characteristic | N (%) or Value |
| --- | --- |
| TOLAC Cohort | 477,693 |
| VBAC Success (%) | 350,340 (73.3) |
| Failed TOLAC (%) | 127,353 (26.7) |
| Maternal Age (SD) | 31.3 (5.1) |
| Pre-pregnancy BMI (SD) | 28.1 (6.7) |
| Gestational Age at Delivery (SD) | 39.1 (1.2) |
| **Race and Ethnicity** |  |
| White | 222,027 (46.5) |
| Hispanic | 123,976 (26.0) |
| Black | 80,333 (16.8) |
| Asian | 30,320 (6.3) |
| Multiracial | 11,036 (2.3) |
| American Indian or Alaska Native | 3,128 (0.7) |
| Native Hawaiian or Other Pacific Islander | 2,021 (0.4) |
| **Number of Prior Cesarean Deliveries** |  |
| 1 | 426,904 (89.4) |
| 2 | 42,464 (8.9) |
| ≥3 | 7,611 (1.6) |
| **Risk Factors** |  |
| Induction of Labor | 141,162 (29.6) |
| Diabetes Mellitus | 46,456 (9.7) |
| Pregestational Diabetes | 5,703 (1.2) |
| Gestational Diabetes | 40,753 (8.5) |
| Hypertensive Disorder | 46,956 (9.8) |
| Chronic Hypertension | 12,668 (2.7) |
| Gestational hypertension | 33,682 (7.0) |
| Eclampsia | 773 (0.2) |
| Prior vaginal delivery | 216,863 (45.4) |
| Maternal age ≥40, N (%) | 24,755 (5.2) |
| BMI ≥30 kg/m<sup>2</sup>, N (%) | 152,706 (32.0) |
| BMI ≥40 kg/m<sup>2</sup>, N (%) | 27,471 (5.8) |
| GA ≥41, N (%) | 56,847 (11.9) |
| **Outcomes** |  |
| NICU Admission Rate (per 1,000) | 58.4 |

#### Cumulative Score Performance

In the national cohort, VBAC success declined as the cumulative risk score increased. VBAC rates ranged from 90.5% at a score of −1 to 44.8% at scores of 4 or higher, with intermediate scores between these values. NICU admission worsened with higher scores, increasing from 43.8 to 111.1 per 1,000 across the same score range, indicating that higher scores identified patients who were less likely to have a successful VBAC and more likely to have an infant admitted to the NICU (Figure 3).

**Figure 3.**
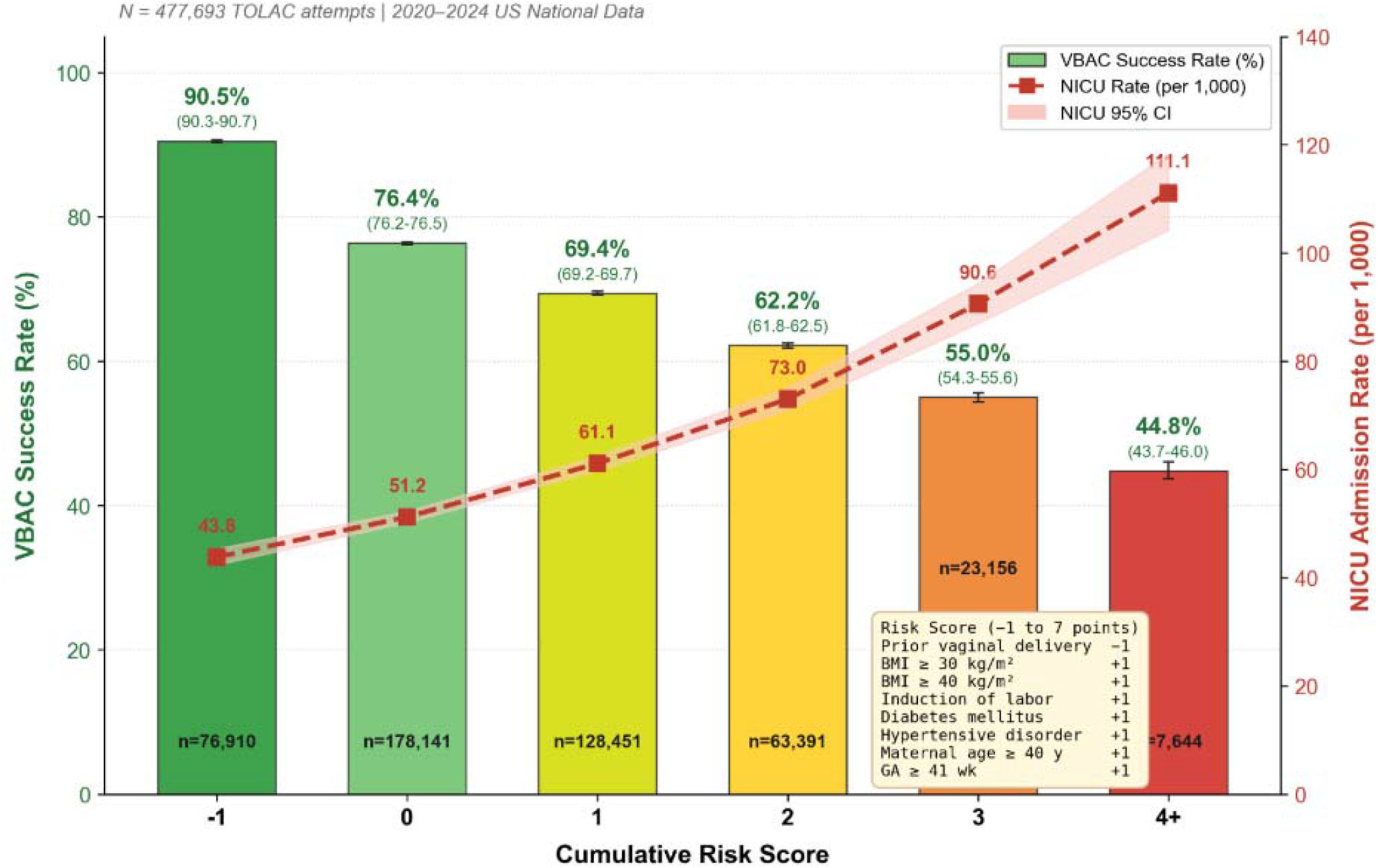
VBAC Success and NICU Admission by Cumulative Risk Score in the National TOLAC Cohort

#### Individual Risk Factor Associations

Prior vaginal delivery was the strongest individual predictor of VBAC success in the national cohort, with VBAC in 86.4% of patients with a prior vaginal birth compared with 62.5% of those without a prior vaginal delivery. Patients with hypertensive disorders were less likely to have a successful VBAC (63.9% vs 74.4% without hypertension). VBAC success was also lower among patients with BMI 40 or greater (58.6% vs 74.2%) and BMI 30 or greater (66.8% vs 76.4%). Diabetes mellitus (67.4% vs 74.0%) and induction of labor (70.8% vs 74.4%) were associated with modest reductions in VBAC success. In contrast, maternal age 40 years or older (70.7% vs 73.5%) and gestational age 41 weeks or greater (72.5% vs 73.4%) were associated with only modest differences in VBAC success.

#### Model Discrimination

In the national validation cohort, logistic regression on a 10% subsample (n=46,598) yielded an AUC of 0.57 for prediction of VBAC success. This discrimination was lower than in the single-center derivation cohort (AUC 0.70) but aligned with expectations for a model derived from clinically detailed institutional data and applied to a national administrative dataset that does not capture variables such as chorioamnionitis, indication for prior cesarean delivery, or Bishop score.

#### Temporal Trends

From 2020 through 2024, the absolute number of TOLAC attempts increased from 89,615 to 101,551; a 13% increase over the study period. National VBAC rates among patients undergoing TOLAC remained stable, ranging from 73.1% to 73.7%, and NICU admission rates remained between 56.5 and 59.8 per 1,000.

## DISCUSSION

In this two-stage study, we developed and externally validated an antepartum cumulative risk score that stratifies both VBAC success and neonatal outcomes. By summing seven routinely available parameters into a −1 to 7-point score, we found that higher scores were associated with lower VBAC probability and higher NICU admission rates. Thus, patients at higher scores are not only less likely to achieve VBAC but also face greater neonatal morbidity. This dual-outcome structure offers a pragmatic tool for risk stratification and shared decision-making by aligning counseling with both VBAC probability and neonatal risk.

Prior vaginal delivery was the strongest predictor of VBAC success. Patients with a prior vaginal birth achieved substantially higher VBAC success than those without, supporting its inclusion as a protective (−1 point) element in the score and highlighting these patients as especially favorable TOLAC candidates. Among the risk-increasing components, hypertensive disorders and severe obesity (BMI ≥ 40) were associated with the largest absolute reductions in VBAC success, followed by BMI 30 or greater, diabetes mellitus, induction of labor, maternal age 40 years or older, and gestational age 41 weeks or greater. Collectively, these patterns show that the score captures a graded spectrum of VBAC likelihood and neonatal risk that can guide TOLAC counseling.

These findings extend prior VBAC prediction work, including the widely used MFMU Grobman model. Existing calculators were developed in earlier cohorts with lower prevalence of obesity and medical comorbidity, require more complex variable sets, and focus on VBAC probability without providing parallel estimates of neonatal risk. In contrast, this dual-outcome score relies on a small set of routinely collected comorbidities, obstetric history, and management decisions available at counseling and links each score level to both VBAC probability and NICU admission, allowing counseling to address maternal mode-of-delivery goals and neonatal risk within a single, simple framework.

The pattern in which higher risk scores concentrate both failed TOLAC and neonatal morbidity supports using this cumulative risk measure not as a gatekeeper but as a structured way to identify patients for whom elective repeat cesarean should be actively considered rather than reflexively dismissed. This score is intended to support individualized counseling aligned with patient preferences and risk tolerance, rather than to replace shared decision making.

An important counterpoint to this precision is that uterine rupture remained unpredictable and occurred at rates comparable to or higher than in some prior cohorts. No individual variable or score component identified a subgroup at meaningfully lower or higher rupture risk, underscoring that rupture is an inherent risk and that TOLAC should be offered only where immediate surgical capability is available.

These findings should be interpreted in light of limitations. The derivation cohort reflects practice at a single tertiary center and lacked some historical details, whereas the national validation relied on birth certificate data that do not capture key variables such as indication and number of prior cesarean deliveries, chorioamnionitis, Bishop score, intrapartum management, or uterine rupture and may misclassify some TOLAC attempts. Differences in NICU admission thresholds across hospitals likely contributed to smaller absolute gradients in neonatal morbidity nationally than in the single-center cohort, and the modest AUC in the national data reflects the expected attenuation when a clinically detailed model is applied to administrative records.

Despite these constraints, applying the same cumulative score to a contemporary national TOLAC population demonstrated reproducible gradients in both VBAC success and NICU admission, suggesting that the tool generalizes beyond a single institution and remains informative in heterogeneous practice settings. Integrating this dual-outcome score into electronic health record calculators and standardized counseling workflows could provide a pragmatic alternative to existing VBAC tools and support more transparent, patient-centered discussions about maternal and neonatal risks and benefits, while helping expand access to TOLAC for patients with favorable profiles and recognizing those for whom elective repeat cesarean may be more appropriate.

## Data Availability

All data produced in the present study are available upon reasonable request to the authors

